# Proteogenomic origins of disease in British South Asians

**DOI:** 10.64898/2026.07.07.26357222

**Authors:** Alice Williamson, Julia Carrasco Zanini, Martijn Zoodsma, Mine Koprulu, Arslan Zaidi, Karen A Hunt, Emma Sol Taylor-Brill, Leonhard Kohleick, Genes & Health Industry Consortium, Genes & Health Research Team, Patrick F Chinnery, William Newman, Sarah Finer, Maik Pietzner, David A van Heel, Claudia Langenberg

## Abstract

Proteogenomic studies have transformed the way we derive novel insights into human biology and pathophysiology but are currently limited by their proteomic coverage and ancestral representation. Here, we integrate rare and common genetic variation with measurements of >11,000 plasma proteins on two affinity-based platforms (>6,000 targets not previously covered) in 1,535 individuals of British Bangladeshi and Pakistani ancestry of the Genes & Health cohort. We report 3,826 high-confidence common (minor allele frequency (MAF)>1.0%) protein quantitative trait loci (pQTLs), over half of which are novel and including >200 pQTLs with greater MAF in South Asians. Systematic analyses of rare (MAF<1.0%) exonic variants identify 230 gene-protein pairs and highlight the joint and distinct contributions of rare and common variants to inherited differences in protein levels. We expand analyses beyond the nuclear genome and identify 3 mitochondrial pQTLs, including a common variant in *MT-RNR1*, associated with lower myelin protein zero (MPZ), identifying a potential novel mechanistic link for *MT-RNR1*’s poorly understood role in hearing loss. We create the first proteogenomic disease network in individuals of South Asian ancestry based on 384 cis-pQTL with a shared genetic disease or risk factor signal, including conditions substantially more common in South Asians, such as metabolic diseases or pregnancy-related conditions, providing insights into the underlying mechanisms. In summary, our study demonstrates the value and scientific efficiency of proteomic studies in genetically informative and understudied populations for identifying novel causes of globally relevant diseases.

## Introduction

Large-scale proteogenomic studies that identify the impact of genetic variation on plasma protein levels and disease have been instrumental in identifying both novel disease mechanisms and candidate targets for pharmacological intervention. Recent successes such as the UK Biobank Pharma Proteomics Project (UKB-PPP) including >50,000 individuals^1–3^ have highlighted the saturation of common cis- pQTL discovery for proteins measured so far in individuals of European ancestry. This, together with the dramatic underrepresentation of non-European ancestries in genomic and proteomic studies^4^ strongly suggest a focus on genetically informative and understudied ancestries, such as South Asian populations, in whom high rates of autozygosity enable discovery of rare homozygous (autozygous) functional variants. In addition, use of multiple proteomic technologies is essential not only for broadest proteomic coverage, but orthogonal validation of pQTLs for proteins with low correlation across platforms.^3,5,6^

Here we present a comprehensive proteogenomic analysis of 1,535 exome-sequenced British Bangladeshi and British Pakistani Genes & Health volunteers, representing ancestries historically understudied in biomedical research.^7–9^ We use the latest versions of two affinity-based proteomics platforms providing extremely broad coverage by targeting 11,327 unique proteins in plasma to study a) rare and common variants across the allele frequency spectrum, b) non-additive modes of inheritance, c) the coding and non-coding nuclear, as well as d) the mitochondrial genome. We highlight 384 protein-disease signals identified through integration of cis-pQTLs with longitudinal health care record derived clinical measures and diagnoses. These protein-disease links advance biological understanding of mechanisms leading to diseases that disproportionally impact South Asian populations, such as type 2 diabetes and obesity. Through integration of variation in the mitochondrial genome, we provide insight into a potential mechanism underlying genetic susceptibility to antibiotic induced hearing loss.

## Results

### Plasma proteomic profiling and genome-wide analyses in Genes & Health

Proteomics profiling of 11,327 unique proteins encoded by 11,341 genes was conducted in up to 1,535 individuals of Bangladeshi or Pakistani ancestry in Genes & Health, using Olink® Explore HT (Olink HT; 5,420 assays, 5,416 unique proteins, N individuals = 1,411), and Somalogic SomaScan® 11K Assay (Somalogic 11k; 11,082 assays, 9,685 unique proteins; N individuals = 1,535) (**Fig. 1a**). This included 6,663 proteins not assessed in pQTL studies to date^1,3,10–23^ and 3,746 captured across both platforms, whose technical performance has recently been benchmarked against mass spectrometry (MS)-based proteomics.^6^

**Figure 1.**
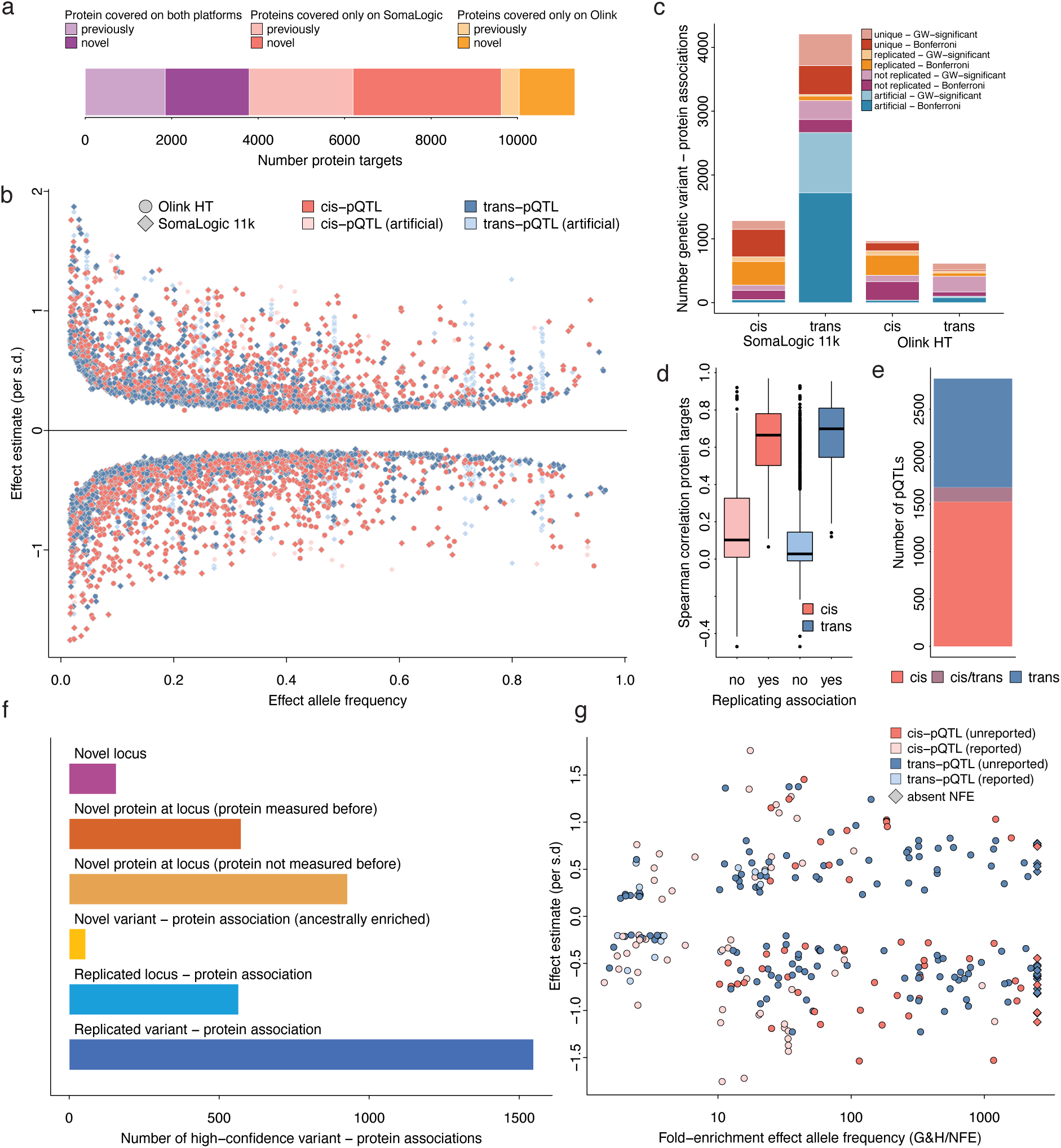
Common variant pQTLs across two proteomic platforms. **a** Fraction of proteins covered with either of the two platforms used (SomaLogic 11k and Olink HT). Darker shades indicate proteins not captured in the so far largest pQTL studies. **b** Scatter plot opposing effect allele frequency and effect size for 7,085 genetic variant – protein associations across both platforms. The colour gradient corresponds to whether the identified variant is close (cis-pQTL) or distal (trans-pQTL) to the protein coding gene. Darker colours indicate high-confidence associations as detailed in **c**. Shapes indicate the platform for pQTL discovery. **c** Distribution of genetic variant – protein associations according to different tiers of confidence and statistical significance (colour gradient). (Bonferroni: p<4.5x10^-12^ for SomaLogic 11k and p<9.2x10^-12^ for Olink HT; GW-significant < 5x10^-8^). **d** Distribution of correlation coefficients for proteins measured on both platforms that were associated with at least one genetic variant stratified by whether the genetic variant was consistently associated across both platforms. Colours indicate the location of the associated genetic variant with respect to the protein coding gene. **e** Number of pQTLs following LD-based clumping of 3826 high-confidence associations across protein targets. **f** Classification of 3826 high-confidence genetic variant – protein associations according to different tiers of novelty. **g** Scatterplot opposing fold-enrichment and effect sizes for 254 variant – protein association with evidence for strongly enriched effect allele frequencies in Genes & Health compared to non-Finnish European ancestry (NFE) subset in gnomAD. Variants that have not yet been seen in NFE are indicated by diamonds.

**Figure 2.**
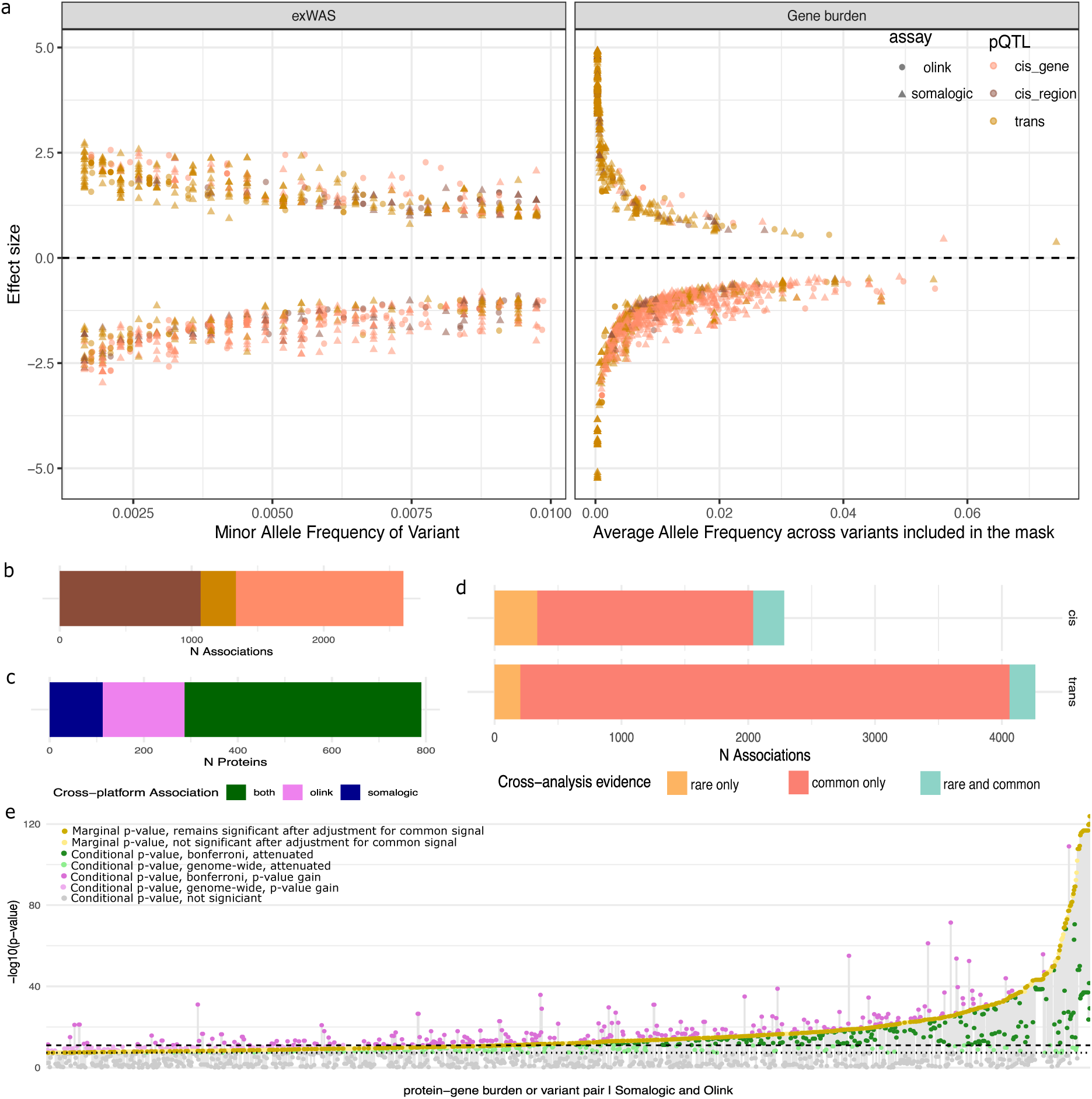
Rare pQTLs across two proteomics platforms. **A**. Identified significant associations across both platforms for single WES variants and gene burden analyses. The x-axis specifies the allele frequency of the variant (exWAS) or average allele frequency of included sites (gene burden), and the y-axis gives the effect size of association. **B**. Number of cis and trans associations identified across the rare variant analyses. **C**. N unique proteins (UniProt ID) with a significant association and their coverage across Olink and Somalogic Platforms. **D**. Comparison of overlap of association evidence for a given gene-protein target pair for cis and trans associations across rare and common variant analyses. **E.** Conditional analyses of rare cis-associations, conditioning for common cis-pQTL signals. Yellow points highlight the marginal p value for association for a given variant/burden-protein target pair. Green points indicate conditional p-value after adjustment for common signal where the p-value drops in significance, and vice versa for pink, where adjustment for common signals increases the significance of the rare association. Dotted lines indicate P<5e-8 and Bonferroni adjusted threshold. Light brown, pink and green, indicate the points that drop to genome wide significance after adjustment, Grey points indicate associations that are attenuated after adjustment for common signals.

#### Common variant regulation (MAF >1%) of plasma protein abundance in individuals of South Asian ancestry

Measurement of plasma protein levels by the latest assay version of the two broadest capture affinity-based technologies (Somalogic 11k and Olink HT) in Genes and Health enabled us to test genetic associations across the allele frequency spectrum targeting over half of the ∼20,000 human protein-coding genes. We identified 7,085 independent genetic variant – protein associations at genome-wide significance (p < 5 x 10^-8^) across the platforms, of which 62.3% (4,421/7,085 associations) also met a more stringent significance threshold adjusted for the number of proteins tested per platform (Olink HT, p < 9.2 x 10^-12^; Somalogic 11k, p < 4.5 x 10^-12^). Overall, we detected 1754 signals in cis (39.6%, Olink: 765; Somalogic: 989 signals) and 2,667 trans signals (60.3%, Olink: 217; Somalogic: 2,450) **(Fig 1b**, Supplementary Table 1).

We systematically screened associations for those likely related to technical factors and artefacts, building on a conceptual framework we recently developed.^5^ Through this process, we identified and subsequently deprioritised 2,853 variant – protein associations (**Fig. 1c**, SomaLogic: n_cis_=48, n_trans_=2665; Olink: n_cis_=36, n_trans_=99) driven by 108 independent (r^2^>0.6) variants associated with one or more of 2504 protein targets (2568 SomaLogic assays, 129 Olink assays). These associations (overlap exists) were a) enriched for proteins strongly affected by technical variation (n=23 pQTL, **Supplementary Table 3**), b) included aptamers targeting non-human proteins (n=38 pQTL), and c) included proteins with genotype-dependent or -induced low correlation across platforms indicative of cis-pQTL epitope artefacts ^5,6^ (n=55 pQTL). We observed moderate correspondence of pQTLs for proteins measured with both platforms (**Supplementary Note**, Spearman’s *r*: cis-pQTL = 0.35; trans-pQTL = 0.09; **Fig. 1d, Supplementary Figure 1**)

To identify a unique set of variants consistently associated across proteomic platforms, we consolidated our finding using linkage disequilibrium (LD: r^2^>0.6) clumping. This identified a high-confidence set of 3,826 variant-protein pairs reflective of 2,823 independent variants that were each associated with at least one protein target across platforms (range: 1-36; **Supplementary table 2**). Most of these pQTLs fell into cis-regulatory regions of the genome, likely acting on the expression or translation of the most proximal protein product (59,3%, n=1,676 / 2,823; **Fig. 1e**). This included 151 cis-pQTLs associated with more than one distinct protein in cis and/or trans. For example, we identified the 3’-UTR variant chr1:109274968:G>T to be positively associated in cis- with plasma levels of sortilin 1 (SORT 1; beta=0.28, p-value<3.9x10^-12^) and cadherin EGF LAG seven-pass G-type receptor 2 (CELSR2; beta=0.37,p-value<6.4x10^-20^) along with 12 proteins in trans. The locus is associated with cardiovascular disease and early work experimentally validated *SORT1* as the underlying causal gene via its role in cholesterol metabolism.^24^ While increasing proteome coverage across platforms leads to broader and more robust discovery, our findings highlight the challenge of leveraging cis-pQTLs that implicate >1 cis-protein to support protein-disease links.

A total of 43% (n = 1,658 / 3,826) of the retained high-confidence genetic variant – protein associations have not been reported so far (**Fig. 1f**), including 132 genetic loci never previously linked to any plasma protein. For example, chr15:85692870:G>A is a common (MAF=45.5%) intronic variant in *AKAP13* is associated with lower plasma levels of the encoded protein, that was not covered in previous proteogenomic studies (beta=-0.77 s.d. units, p-value<3.6x10^-90^). AKAP13 is a guanine nucleotide exchange factor that activates the small GTPase Ras family homolog member A (RhoA).^25^ Variants in strong LD (r^2^>0.8) at this locus have previously been reported to be associated with shifts in white blood cell composition and increased blood pressure^26^, and we provide human genetic evidence for the involvement of AKAP13 in blood pressure regulation.^27,28^ A role of AKAP13 in sensing osmotic stress may explain its dual effect on immune cell populations and blood pressure, with high salt intake being among the most common risk factors for high blood pressure and AKAP13 promoting the expression of cytokines and subsequent differentiation of B cells in lymphoid organs.^29^

Expansion of proteomic coverage of the latest assays contributed to most of the novel signals (56,8%, n=942 / 1,658), including 237 of the identified cis associations. These cis associations, in some cases, provided valuable context for trans associations at the same locus. For example, the chr12:89458668:C>T (MAF=21.9%) has repeatedly been reported as a pleiotropic trans-pQTL.^1–3,12,14,20,21,30^ We identified, for the first time, its cis association with higher plasma levels of polypeptide N-acetylgalactosaminyltransferase 4 (GALNT4; beta=0.70, p-value<4.3x10^-62^). GALNT4 catalyses the initial step in O-linked oligosaccharide biosynthesis, which represent an important posttranslational modification for a broad spectrum of protein targets, thereby identifying the likely reason for the pleiotropic trans associations seen at this locus.^31^

### Variants with increased frequency contribute to the risk of diseases more prevalent in South Asians

Differences in allele frequency are a good indicator of true differences in genetic associations across ancestries.^2,32,33^ We identified 254 variant – protein associations that showed strong evidence of ancestral enrichment in our study (driven by 90 independent cis and 127 trans variants), with higher minor allele frequency in South Asian populations compared to Non-Finnish Europeans (NFE). We defined ancestral enrichment as a >25% increase in allele frequency or >10-fold difference compared to European studies (**Fig. 1g**). Of the 90 cis-pQTLs, 52 variants (57.7%) are unreported in previous, mostly European-based pQTL studies. This included twelve proteins, five of which not previously assayed, that are targets of drugs currently approved or under active development. For example, the missense variant chr18:63318540:C>T (rs1800477, p.Ala43Thr) within *BCL2* is common in our study population (MAF=7.65%), but rare outside of Asia (e.g., MAF=0.02% in NFE). We identified an inverse association with plasma levels of the protein product BCL2, an apoptosis suppressor (BCL2; beta = -0.87, p-value<5.1x10^-39^). BCL2 is critically involved in the pathogenesis of cancers, including chronic lymphocytic leukaemia (CLL)^34^ for which the BCL2-inhibitor venetoclax was approved in 2016.^35^ *BCL2* p.Ala43Thr has been functionally characterised *in vitro* to impact levels of BCL2, consistent with our observation, and further increased sensitivity of cells harbouring this variant to chemotherapy-induced cytotoxicity and apoptosis compared to those expressing wild type BCL2.^36,37^ The prevalence of CLL is rare in Asian compared to Western countries^38,39^ and while the exact mechanism is not understood, a genetic contribution through ‘protective’ variants, such as suggested for p.Ala43Thr, is plausible. The *BCL2* locus has been associated with lower risk of CLL in European populations ^40,41^ based on a distinct common variant (rs4987845, MAF_EUR_=9.3%; LD_SAS_=0.003; D’_SAS_=1.0; LD_EUR_=0) upstream of *BCL2* that is found at low frequency in South Asian populations (MAF_SAS_=2.2%). This variant (rs4987845) has not been associated with plasma BCL2 in much larger studies but is associated with BAFF receptor levels^2^, an activating factor of disease-causing B-cells. This illustrates the value of ancestrally diverse pQTL studies to guide identification of ethnic-specific protective or risk mechanisms for disease.

A further example included chr4:38797027:C>A (rs5743618;p.Ser602Ile), a cis-pQTL for Toll-like receptor 1 (TLR1) (beta = 0.51, se=0.06, p = 2.51x10^-15^) that is found at different frequencies across ancestral groups (MAF_SAS_: 91%; MAF_NFE_: 27%) and suggested to be under strong positive selection.^42–44^ TLR1 is a pattern recognition receptor from the innate immune system, forms a heterodimer with TLR2 and activates the innate immune system upon detection of microbial membrane components.^45^ TLR1 p.Ser602Ile has been reported to result in lower TLR1/TLR2 heterodimer expression on the cell surface of immune cells and reduced TNF-alpha activation, indicative of reduced NF-kB pathway activation.^46–48^ Large scale GWAS have associated the effect allele (A-allele) with lower risk of asthma, allergic rhinitis and pollen allergies, in line with a lower risk of hyperresponsiveness of the immune system.^49–51^

### Non-additive effects on the proteome

Enrichment of homozygous non-reference alleles in Genes & Health participants^52^ provided us with the unique opportunity to test for the presence of non-additive effects. We identified evidence of non-additive effects for a total of 65/3,826 pQTLs identified in our primary additive analyses (**Supplementary Table 4**). For example, the cis-pQTL chr14:20472447:G>A for plasma purine nucleotide phosphorylase (PNP) levels (rs1049564; p.Gly51Ser) was best explained under the recessive model (beta = -1.17, p = 2.12 x 10^-24^) compared to the additive models originally used for initial discovery (beta = -0.34, p-value=6.12x10^-10^) (**Supplementary Fig. 2**). Patients homozygous or compound heterozygous for damaging mutations in PNP have nucleoside phosphorylase deficiency (OMIM 164050; ^53–55^), resulting in severe combined immunodeficiency due to defects in T-cell function and/or levels. Furthermore, rs1049564 has been shown to result in reduced PNP protein and mRNA levels and has also been reported to be associated with systemic lupus erythematosus, further suggesting a role of PNP in immune-related conditions.^56^

### Rare exonic variant contributions to the plasma proteome

We next expanded our analyses to protein association of rare coding variants (MAF < 1%) captured by whole-exome sequencing, identifying a total of 1020 rare variant – protein associations, of which 23.4% (239/1020 associations) passed a more stringent Bonferroni significance threshold, including 175 unique protein targets and 190 unique genetic variants. (**Supplementary Table 5 and 6**). We identified pronounced allele frequency differences compared to European populations for these rare, predominantly coding variants, including 36/190 variants (28% of associations) absent in NFE populations according to the gnomAD Exomes or Genomes database^57^, but also variants that were ultra-rare in NFE populations (MAF < 1 x 10^-5^). For example, we identified a rare coding cis-pQTL predicted to be damaging (CADD = 24.9, DDG=3.03^58^) and associated with lower calcium-binding and coiled-coil domain 2 (CALCOCO2) levels (chr17:48848379_G>C, p.Gly114Ala, beta =-1.56, p-value < 2.1 × 10⁻¹⁵). The variant was found in 0.7% of G&H participants and 0.4% of South Asians in gnomAD, but only a single heterozygote out of 1.2 million gnomAD NFE samples. CALCOCO2 has been identified to be a regulator of pancreatic beta cell function^59^, a key pathway implicated in T2D development and specifically the early onset of T2D in South Asian populations compared to European populations.^60,61^

To increase power for rare variant analyses, we tested aggregated burden effects and identified evidence for 232 gene burden mask-protein associations, including 187 unique gene-protein pairs at Bonferroni significance, of which 145 acted in cis. Around 70% of the proteins identified in gene burden analyses also had evidence of rare single variant effects (above). The genetic background in this population and expanded proteomic coverage of the Olink platform allowed us to i) identify cis-associations for proteins unseen in UKB-PPP despite a ∼45x larger sample size (4 cis-associations for DIPK2B, FCGR2A, HJV and IFI30), and ii) identify cis-associations for proteins not covered in the UKB-PPP (6 cis-associations for ACP*3, ECI1, FTO, IAH1, RIGI, SAA4*).

### Independent rare and common variant effects

To identify whether nearby common and rare genetic signals associated to the same protein were independent of each other, we performed conditional analyses for 260 rare cis associations significant at P < 5x10^-8^ (219 unique variants and 150 unique proteins) with evidence of a common nearby cis-pQTL signal (+/-500kb. Common cis-pQTL P < 5e-8). After conditioning on common nearby cis-pQTL signals, 186 of these 260 associations remained significant (P < 5x10^-8^), 67% (67/100) for Olink, and 74.3% (119/160) for Somalogic (**Supplementary Table 7**).

For 373 of the 643 cis associations identified through gene burden tests reflecting unique single gene mask-protein target pairs, there was evidence of a nearby common cis-pQTL. These reflected 219 gene-protein target pairs implicating a minimum of one mask per gene, of which 213 remained significant after adjustment for nearby common cis-pQTL signals, 93% (82/88) for Olink and 100% (131/131) for Somalogic at P < 5x10^-8^ (**Supplementary Table 9**) in conditional analyses.

Together, this highlights the distinctness of rare and common variation in cis contributing to plasma protein levels, in line with studies of other human traits ^62,63^.

### Mitochondrial DNA analyses highlight interactions between the mitochondrial genome and circulating levels of nuclear-encoded proteins

Variation in the mitochondrial genome and mitochondrial dysfunction is known to influence risk of common complex diseases including type 2 diabetes and cardiomyopathy.^64–68^ We identified three distinct significant associations between homoplasmic mitochondrial single nucleotide variants (mtSNVs; MAC > 5) and plasma levels of proteins encoded in the nuclear genome (p<5x10^-8^; **Supplementary Table 10**). To our knowledge this is the first mitochondrial genome pGWAS studies to date.

Nuclear-encoded proteins associated to mtSNVs were directly implicated in mitochondrial function. We did not identify evidence for rare or common pQTLs in the nuclear genome for two of the three significantly associated proteins and observed that the estimated effects of mtSNVs on these proteins was much larger compared to similarly frequent nuclear-encoded pQTLs (**Fig. 3b**). For example, the mtSNV MT:980:T>C (rs397515731) was associated with higher plasma levels of the nuclear encoded mitochondrial ribosomal protein S11 (MRPS11; beta = 0.38, p-value = 2.14 x 10^-9^), which encodes the small 28S subunit protein in mitochondrial ribosomes. MT:980:T>C (rs397515731) is located within *MT-RNR1*, which encodes the mitochondrial 12S rRNA, which directly interacts with the small 28S subunit, and therefore is directly implicated in a protein complex within the mitochondria.^69^

**Figure 3:**
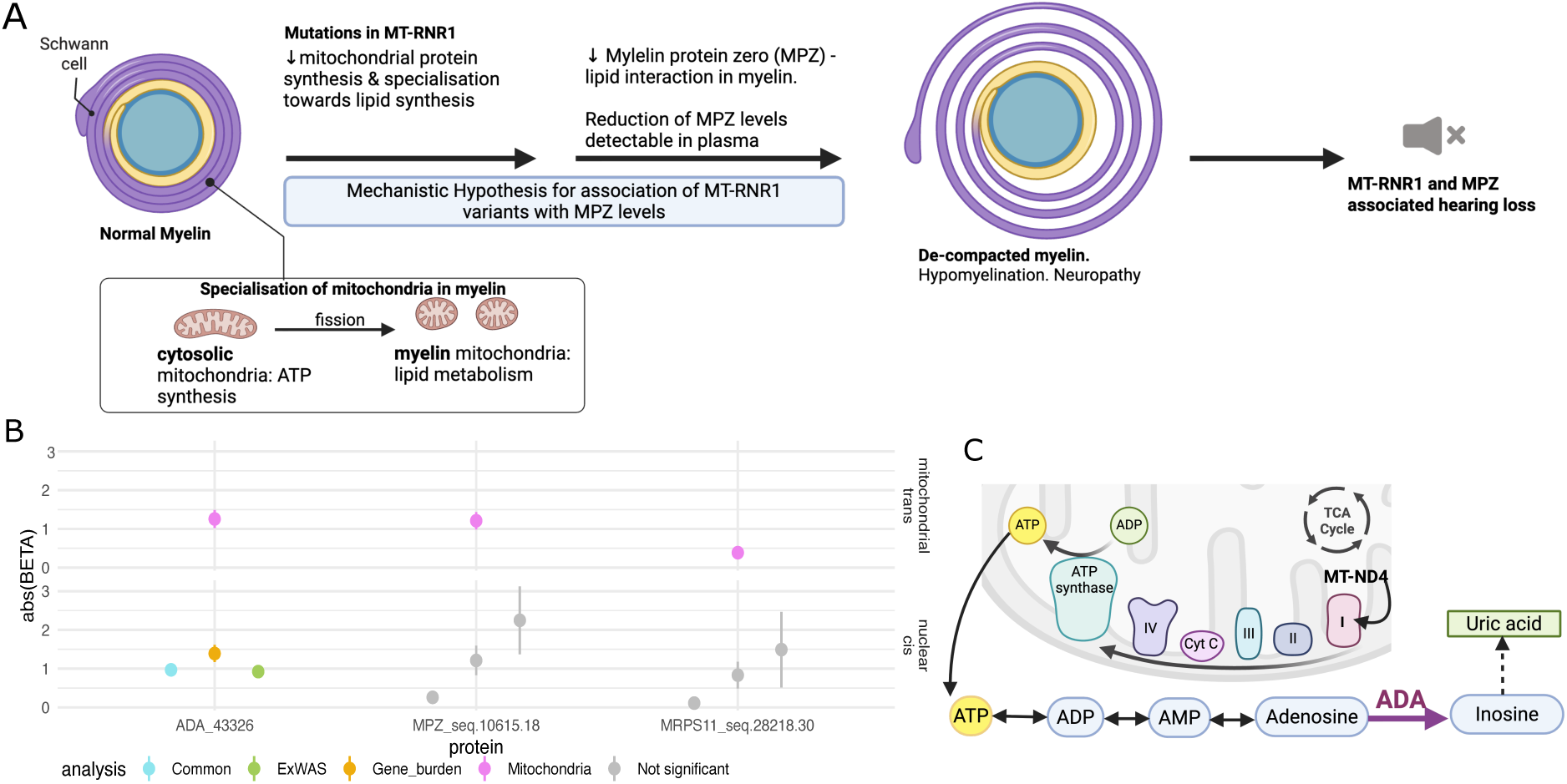
Mitochondrial Genome wide association study of the plasma proteome for proteins measures on either Olink or Somalogic. **A**) Hypothesis of candidate mechanism of action linking variants within MT-RNR1 to reduced MPZ levels and subsequent links to hearing loss. **B**) Effect size plot for the 3 identified significant mitochondrial genome associations and cis-pQTLs for the protein target in the nuclear encoded genome (rare – exWAS and gene burden and common – GWAS). Point represents the absolute effect size and error bars +/- standard error. Significance of the result is highlighted by opacity of the points. For gene burden and exWAS associations the most significant association by p-value was represented on the plot. The cis-PQTL for the common GWAS is the most significant associated variant within +/-500kb of the gene body as previously described. **C**) Candidate mechanism linking variants within MT-ND4 to reduced levels of ADA in plasma.

We further identified MT:1018:G>A (rs2856982) within *MT-RNR1* to be associated with reduced plasma levels of myelin protein zero (MPZ; beta = -1.21, se = 0.22, p-value = 4.79 x 10^-8^, EAF=0.0026). MPZ is the most abundant protein in myelin and is specifically expressed in Schwann cells of the peripheral nervous system.^70^ Rare damaging mutations within *MPZ* are known to cause neuropathies including in Charcot-Marie-Tooth, Dejerine-Sottas disease, hypomyelinating neuropathy and Roussy-Levy syndrome.^71,72^

In vivo studies have demonstrated that *MPZ* mutations cause neuropathy via different mechanisms resulting in hypo- and demyelination. This includes through myelin instability resulting in defects in packing of the myelin sheath and mutations resulting in initiation of the unfolded protein response resulting in lack of or insufficient myelination (**Fig. 3a**)^73^ These different pathologies have also been observed through light and electron microscopy characterisation of nerve biopsies of a cohort of patients with *MPZ* mutations when comparing to controls without *MPZ* mutations.^74^

Finally, MT:11128:A>G (rs1603223120) within *MT-ND4* was associated with reduced adenosine deaminase (ADA) abundance in plasma (beta = -1.26, p-value < 4.77 x 10^-8^), a key enzyme in purine metabolism. MT-ND4 encodes NADH dehydrogenase 4, which forms part of complex I in the mitochondria, an essential protein complex in the electron transfer chain in ATP synthesis (**Fig. 3c**). Adenosine, which is converted to inosine by ADA, is derived from the process of ATP breakdown. Therefore, dysfunction in this complex due to variation could result in reduced ATP synthesis and therefore lower substrate, meaning lower ADA enzymatic activity is required during purine metabolism.^75^

### A proteogenomic disease network in individuals of South Asian ancestry

We performed systematic genetic colocalisation analyses to identify genetic associations shared between the proteome and phenome. Genetic colocalisation analyses highlight whether genetic signals observed at a given locus for two traits, in this case a protein and a disease/related trait are shared and likely driven by the same causal variant. We integrated evidence for shared signals from gene to protein to disease for all common (MAF >1%) cis-pQTLs (or their proxies, R^2^>0.7) associated with diseases or traits at P<1e-4. We used multi-source electronic health records available for >43,000 G&H volunteers with genetic information, specifically a) 3-digit ICD10 codes, b) G&H curated disease definitions (see methods), and c) EHR-derived clinical measures. We identified a total of 384 unique connections (posterior probability of a shared signal (PP) >0.7), implicating 229 unique proteins (288 protein targets) and 228 partially overlapping disease definitions (**Fig. 4 and Supplementary Table 11**). A further 179 links were identified between cis-pQTLs and clinical measurements, including blood tests, spanning 74 proteins (99 protein targets) and 38 clinical measures.

**Figure 4:**
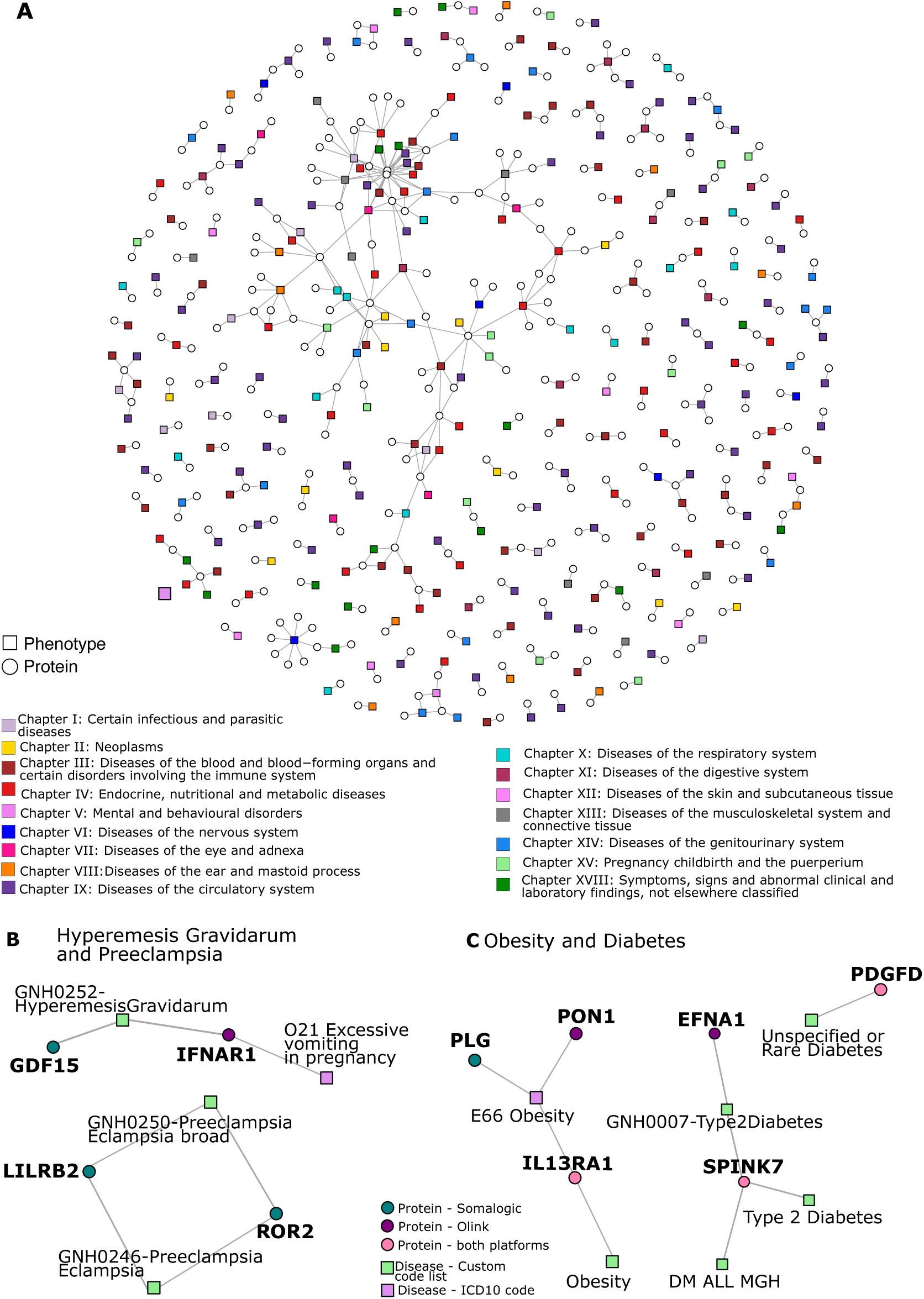
Protein disease links identified through cis-pQTL colocalisation. **A**) Protein-disease network showing protein-disease links where there was evidence of a significant colocalisation between a cis-pQTL and disease/trait (PP.h4 > 0.7, Rsq lead variants >= 0.7). Colours represent the ICD10 chapter to which diseases/traits were mapped. **B**) cis-protein-disease links for pregnancy related conditions Hyperemesis Gravidarum and preeclampsia **C**) cis-protein-disease links for obesity and diabetes. Circles represent the cis-proteins with colours representing the platform for which the given protein target had evidence of colocalisation. Squares represent disease outcomes with colours representing the source of the disease definition.

These included 80 connections at previously unreported cis-loci (n=45 unique proteins) and a further 232 connections (n=146 unique proteins) for which the cis-pQTL or proxies (r^2^>0.8) had not been reported for any non-proteomic trait in the GWAS Catalog. This highlights the value of systematically integrating clinical data from health records with expanded protein coverage in a genetically informative and underrepresented ancestral group.

Most identified connections related to the blood and immune system (83 protein-disease pairs, 14.7%), diseases of the circulation (100 protein-disease pairs, 17.8%), or endocrine and metabolic diseases (79 protein-disease pairs, 14%) (**Fig. 4b**).

While most cis-pQTLs with evidence of a shared genetic signal with a disease showed comparable frequencies across major ancestral groups, we identified two examples with strong enrichment in British South Asians. For example, we observed evidence for the cis-pQTL chr3:3108627:C>T (rs77530409; MAF_SAS_=27.5%, MAF_NFE_=0.6%), associated with higher plasma interleukin 5 receptor alpha subunit (IL5RA measured by Olink,; beta=0.56, p-value=3.16x10^-50^) to be a shared signal for the risk of angina pectoris (ICD10 code I20; beta=0.134, p-value=2.82x10^-4^; PP=76.8%). The cis-pQTL for IL5RA is rare outside of Asia and not in LD (r^2^_max_=0.05 across ancestries) with a common signal previously identified at the locus in Europeans tagged by rs77400868 (MAF_SAS_=9.8%; MAF_NFE_=13.4%).^2,14^ rs77400868 is in perfect D’ (D’=1 across ancestries) with the cis-pQTL identified in this study (rs77530409) suggesting that both alleles share an ancestral haplotype. The ancestral complexity of the locus was further emphasized by the observation that the cis-pQTL for plasma IL5RA levels, as measured by SomaLogic, chr3:3108107:A>G (rs3792421 MAF_SAS_=44.3%; beta =0.58, p =1.58x10^-66^ protein target ID = seq.4491.4), was twice as common in British South Asians compared to Non-Finish Europeans (MAF_NFE_=26.7%), showed evidence of a shared genetic singal for coronary heart disease (PP=88.2%; beta =0.12, p = 1x10^-4^), was in partial LD (r^2^=0.48) with the Olink-associated lead variant but only in D’ with the European lead variant for IL5RA as measured across assay systems (r^2^_max_=0.12, D’=1.0). This discrepancy in identifying different lead cis-pQTL variants across ancestries might explain why the European signal has not been reported to be associated with similar diseases in much larger European cohorts as recorded in the Open Targets portal [download 26/09/2025]. Further, while we provide robust evidence of a shared signal through genetic colocalisation analyses (see above), the genetic association with angina pectoris or coronary heart disease does not meet conventional genome-wide statistical significance and requires replication, particularly in south Asian cohorts. There is no evidence of this locus being associated with coronary heart disease in published studies, which did not include individuals of South Asian Ancestry in large databases such as the Common Metabolic Disease Knowledge Portal and Open Targets^76–78^. However, the evidence from our study in Genes & Health of a shared genetic signal of a cis-pQTL for IL5RA measured by two technologies with two related cardiac conditions, highlights the need for further investigation of this identified gene-protein-disease link.

### Diseases prioritised through engagement of the local East-London community

Both Queen Mary University of London and the Genes & Health team^79^ have engaged the relevant local and study communities, respectively, to identify views on their priority areas for research. These engagement exercises highlighted women’s health, including conditions in pregnancy, and metabolic diseases, among other conditions of high prevalence in South Asian populations. Here, we focus on examples from some of these areas and integrate our findings with orthogonal evidence from the literature and animal models.

#### Pregnancy related conditions

Women of South Asian ancestry are at increased risk of preterm birth and other pregnancy-related conditions.^80^ The mechanisms underlying such conditions are not fully understood. The detailed recording of pregnancy-related outcomes through ICD10 codes and custom code lists in G&H enabled us to identify protein-disease links for important pregnancy-related conditions. We identified evidence of effects of loci for cis-protein levels of 23 proteins to be shared with pregnancy outcomes including conditions occurring at different stages during gestation. For example, we identified a shared genetic signal between a cis-pQTL for IFNAR1 (PP=92%; chr21:33317653:A>T beta=0.36, p = 6.03x10^-12^), and excessive vomiting in pregnancy (beta=0.20, p=1.17x10^-4^). IFNAR1 forms heterodimers with INFAR2 on the cell surface to form a receptor for type 1 interferons and therefore plays a role in activation of the TYK2-STAT signalling cascade.^81^ To our knowledge IFNAR1 has not been linked to this condition previously. In addition, we identify a cis-pQTL of GDF-15 (MIC1) and excessive vomiting in pregnancy and hyperemesis gravidarum (HG; PP=99.9%; chr19:18371548:G>A; GDF15: beta=0.26, p-value=6.17x10^-18^; HG beta =-0.26, p-value = 2.88x10^-11^). While a directionally consistent link for GDF15 (MIC-1) has previously been established in European ancestry individuals,^82,83^ our study provides the first evidence in individuals of South Asian ancestry.

South Asian women are also at higher risk of preeclampsia,^84^ a multi-system disease with hypertension during pregnancy (> 20 weeks gestation) and systemic inflammation posing a significant risk for mother and child.^84^ Preeclampsia is driven by dysfunctional placenta including impaired trophoblast invasion resulting in impaired uterine artery remodelling and angiogenesis. The aetiology is currently incompletely understood but several genetic loci have been identified to be associated with increased preeclampsia risk for the mother in both the maternal and offspring genome.^85–88^ We identified two new gene-protein links for pre-eclampsia (LILRB2, ROR2).

ROR2, is a receptor tyrosine kinase that has been reported to play a role in trophoblast viability and migration.^89,90^ This is to our knowledge the first study to provide human genetic support for the link between ROR2 protein levels and preeclampsia. This supports observations that Placental derived ROR2 has been reported to be elevated in preeclampsia patients, based on samples from the antecubital vein.^91^

LILRB2, belongs to the leukocyte immunoglobulin-like receptor family and is expressed predominantly in innate immune cells where it plays a role in immune inhibition.^92^ LILRB2 is expressed in the placenta and umbilical cord during pregnancy and plays a role in the HLA-G pathway which is important for maintaining immune tolerance at the maternal-foetal interface.^93^ The role of HLA-G and LILRB2 in immunosuppression has been reported to play a key role during placentation.^92^ In preeclampsia it has been demonstrated that LILRB2 and other proteins within this pathway are downregulated in expression in placenta from mothers with preeclampsia compared to mothers without.^94^

#### Metabolic diseases (Diabetes and Obesity)

Metabolic diseases linked to adiposity, such as type 2 diabetes are much more prevalent in South Asian compared to European populations, despite on average lower BMI.^95^ It is established that individuals of South Asian ancestry develop the metabolic complications of obesity at lower BMI levels compared to Europeans, and this is reflected in ancestry specific diagnostic threshold recommended in clinical practice.^96,97^ For example, for assessing T2D risk ancestry specific BMI cutoffs are recommended as 30 kg/m^2^ in Europeans but 23.9 kg/m^2^ in South Asian Populations.^97^ We identified shared genetic cis-signals for 3 proteins (EFNA1, SPINK7, PDGFD) and diabetes risk, with some studies reporting evidence supporting these new links.^98–102^

In addition, we identified a shared genetic architecture for 3 proteins, IL13RA1, PON1, PLG, and the diagnosis of clinically diagnosed obesity, as evident from specific ICD10 codes in health records of G&H participants. These included established links for cytokine IL13RA1, a subunit of the IL13 receptor complex, in obesity and adipose tissue remodelling and fibrosis.^103–105^ IL13 and IL13RA1 have further been recently reported to be involved in regulating beige adipogenesis, the browning of white adipocytes, including a role in modulating thermostasis.^105^ This is of relevance to obesity with browning of adipocytes proposed as a therapeutic avenue for obesity.^106^ We identified evidence for a shared genetic signal for IL13RA1 and obesity across protein targets for IL13RA1 across both Olink and Somalogic, and multiple obesity definitions (e.g. PP = 94%, cis pQTL = chrX:118770222:A>G, Olink IL13RA1 effect = 0.689, SE = 0.033, p-value =1.74x10^-95^; Obesity effect = 0.085, SE = 0.01, p-value = 5.89x10^-7^; **Supplementary Table 11**).

We additionally identify a protein-disease links for obesity with levels of PLG (PLG Somalogic cis-pQTL = chr6:160664088:T>A ; PP=87%, PLG effect = -0.198, se = 0.0355, p-value = 2.36x10^-8^; obesity = -0.0738, se = 0.0193, p-value = 1.34x10^-4^) and PON1 (PON1 Olink cis-pQTL = chr7:95324583:G>A; PP = 94.4%; PON1 Olink effect = -0.4996, se = 0.036, p-value = 1.35x10^-43^; Obesity beta = 0.074, se = 0.018, p-value = 3.47x10^-5^), with direction of effect of the lead cis-pQTL variants on protein levels and obesity directionally consistent with experimental evidence in the literature (**Supplementary Table 11**). PLG, Plasminogen, plays a role in fibrinolysis.^107^ Mice lacking *Plg* have been identified to have reduced adipocyte accumulation and lower body weight, directionally consistent with the human genetic evidence observed.^108^ PON1 has been identified to be involved in HDL cholesterol regulation and regulated by leptin.^109^ It has been further reported to be involved in downstream consequences of obesity such as insulin resistance^110^ and regulation of GLUT4 expression, the primary postprandial glucose transporter in muscle and adipose tissue.^111^

## Discussion

Here we combine two of the most comprehensive affinity-based broad capture proteomics technologies to date with systematic assessment of rare and common genetic variation in a cohort of understudied South Asian ancestry individuals. This allowed us to identify novel proteogenomic links and, through integration with electronic healthcare records, further understand their contribution to disease risk across the phenome. We report novel associations for newly covered protein targets, but also identify novel associations for previously assayed proteins, likely due to differential effect allele frequencies within G&H compared to European studies that have dominated previous efforts. Our study highlights that not only increased breadth of protein coverage by proteomics platforms but also analyses in understudied ancestry groups provide increased discovery.

Building on previous work^6^, we flag possibly artificial common pQTLs that are likely explained by technical artefacts rather than robust genetic associations. We further use converging proteomic measurements across platforms to identify genetic variants most consistently associated, although we note that the concordance between platforms was moderate, as previously reported.^3,5,112,113^ Such analyses highlighted the strength of parallel assessment of the plasma proteome across two platforms to identify the most robust associations.

We note that of the subset of pQTLs found more frequently in individuals of South Asian ancestry compared to most widely studied European ancestry individuals include a cluster of genetic loci centred around immune genes. In line with the literature, these may potentially highlight evolutionary selection of such loci, including in south Asian populations.^114–116^ This emphasises the importance of expanding proteogenomic efforts to diverse ancestry groups and the ability to discover novel proteogenomic associations even in modest sample sizes due to differential frequency of alleles across different populations. We identified allele frequency differences in cis-variants affecting the levels of several proteins including TLR1, AKAP13 and GALNT4 which are highlighted in detail above, all of which are implicated in regulation of nuclear factor-kappaB (NF-κB) activation, a key processes in the innate immune response and inflammation. Toll like receptors (TLRs) are patten recognition receptors that detect microbial components.^117^ NF-κB is a family of master transcription factors that regulate expression of key genes implicated in the immune response. For example: TLR1 forms heterodimers with other TLRs, predominantly TLR2, and this heterodimer recognises lipopeptides on bacteria, activating the downstream immune response. AKAP13 has been demonstrated to play a role in the pathway downstream of TLRs by modulating NF-kB activation.^25^

We study non-additive effects and the contribution of mitochondrial genetic variation provided novel insights into the genetic regulation of circulating protein levels, going beyond additive effects in the nuclear genome that have predominately been studied. These include a possible mechanistic link between *MT-RNR1* variants and hearing loss, even in the absence of aminoglycoside exposure. The associated protein product myelin protein zero (MPZ) warrants further investigation as a novel candidate circulating biomarker for hearing loss and neuropathy. Mitochondria play an important role in myelination, a metabolically intensive process. Mitochondria in myelin are functionally different to cytosolic mitochondria, and play a specialized role in lipid metabolism, suggesting that variation that affects this shift in mitochondrial function within this cellular compartment may impact this process.^118^ Abnormal mitochondrial function and morphology in Schwann cells have been linked to aberrant lipid metabolism within Schwann cells and neuropathy.^119,120^ Some *MT-RNR1* variants have been identified to result in aminoglycoside induced sensorineural hearing loss as these variants cause *MT-RNR1* structure to more closely resemble the prokaryotic 16s rRNA, the target of aminoglycoside antibiotics. Therefore, genetic testing for three clinically well-established variants known to result in increased risk of aminoglycoside-induced hearing loss, which are different to the variant found in the current study (m.1095:T>C - MAF_NFE_ = 0.00027 MAF_SAS_ = 0.00067, m.1494C>,T- MAF_NFE_ = 0.00039, MAF_SAS_ = 0.00067, m.1555A>G - MAF_NFE_ = 0.00097, MAF_SAS_ = 0.0013. Frequencies reported in Gnomad r4 ^121^). Clinical genetic screening programmes for MT-RNR1 variants are now recommended, for example, for patients predisposed to risk of gram-negative infection due to conditions such as cystic fibrosis.^122^ A study in a Han Taiwanese population previously highlighted that variants within *MT-RNR1* are associated with hearing loss even in the absence of aminoglycoside exposure.^123^ MPZ could be hypothesised to be a candidate mediator between MT-RNR1 and hearing loss in absence of aminoglycoside exposure and warrants further investigation. Collating these results, we hypothesise that reduced MPZ levels are a marker of uncompacted myelin (**Fig. 3a**).

Finally, the comprehensive phenome-wide assessment of sharedness of common pQTLs with diverse disease endpoints highlighted several candidate novel, proteogenomically anchored, protein disease links across the phenome, including for diseases that disproportionally impact South Asian populations, such as type 2 diabetes. These analyses also provided in some cases the first proteogenomic evidence for known protein-disease links also in individuals of South Asian ancestry (e.g. GDF15 and hyperemesis gravidarum), contributing to more equitable biomedical and genetic research.

While our discovery sample size is comparatively modest compared to emerging large scale proteogenomic studies, it focuses on a genetically informative population of individuals of South Asian ancestry underrepresented in research with a significant burden of disease and premature mortality. However, while our work addresses an important gap in the literature, future proteogenomic studies more inclusive of other global, ancestrally diverse populations are needed.

As has been described previously, use of electronic healthcare data for disease endpoint identification comes with some limitations reporting and capture of certain conditions. However electronic healthcare data is more robust to challenges seen in traditional population studies as it minimises selective loss of follow up due to lack of ongoing participation. The power to detect disease associations in G&H is limited due to sample size and therefore prevalence of these conditions, we therefore ensured support for protein disease links for the highlighted examples through literature review and evidence from larger genetic studies. We further accounted for possible false positives in our genetic colocalisation analyses through a staged approach filtering by p-value in the disease/trait GWAS, posterior probability and LD between lead variants. However, it is essential that these findings are replicated in future studies to ensure stability of these results.

To conclude, we provide a comprehensive assessment of the plasma proteome using two affinity-based proteomics platforms across the allelic spectrum, inheritance patterns, and nuclear and mitochondrial genome, focussed on underrepresented individuals of Bangladeshi and Pakistani ancestry. Our results not only demonstrate the value of ancestrally diverse proteogenomic studies to identify novel disease mechanisms, which are relevant to inform drug discovery strategies across broader ethnic groups, but also provide a resource that will enable integration of these data with other cohorts of South Asian ancestry. Through integration this omics data can be used to investigate and prioritise candidate causal genes in GWAS studies of other traits and phenotypes.

## Materials and Methods

### Study design

G&H is a cohort of British Bangladeshi and British Pakistani individuals in the United Kingdom (UK).^79^ To date, over 70k individuals have been recruited, with up to 55k being whole-exome sequenced at the time of this analysis, focusing on the subset of ∼1500 recalled for plasma sampling and proteomic analyses, as described below. Inclusion criteria included individuals aged 16 and over, who self-identify as being from Bangladeshi and Pakistani backgrounds. Recruitment has been ongoing since 2015 in healthcare and community settings. A brief demographic questionnaire was performed at recruitment, as well as collection of a saliva sample for genotyping and sequencing, and consent for invitation to recall and to longitudinal electronic health record (EHR) data linkage including data from National Health Service primary and secondary care, Hospital Episode Statistics, and Cancer death registry. Details of the cohort have been previously described. G&H was approved by the London Southeast NRES Committee of the Health Research Authority (14/LO/1240).

The current study comprises a subset of participants from the G&H cohort that were recalled for additional phenotyping and blood sampling. EDTA-plasma samples were collected and processed within ≤1 hour and subsequently stored at -80°C until further analysis.

### Proteomics profiling – Olink and Somalogic

#### Olink Explore HT

Proteomic profiling with the Olink Explore HT platform was performed in 1641 samples from 1,447 individuals. Details of the Olink platform have been previously described in detail.^124^ Briefly, Olink relies on proximity extension assays, which targets proteins by pairs of antibodies conjugated to complimentary oligonucleotides. Protein assays are grouped across eight dilutions blocks, with blocks 1 to 4 having a 1:1 dilution (i.e. including low abundant proteins in plasma), block 5 a 1:10 dilution, block 6 a 1:100 dilution, block 7 a 1:1,000 dilution and block 8 a 1:100,000 dilution (i.e. including the most abundant proteins in plasma). Upon binding to their target protein, hybridization between probes enables amplification and subsequent relative quantification through next generation sequencing. Olink’s internal controls involve an incubation (a non-human antigen with matching antibodies), extension (IgG conjugated with a matching oligo pair) and amplification controls (synthetic double stranded DNA). Additional external controls are included in each plate, namely negative, plate (increased to 5 compared to the 3 included in their previous version of the Explore platform) and sample controls. Normalised protein expression (NPX) values are generated by normalisation to the extension control, log2 transformation and further normalisation to the plate controls. Samples are flagged as a FAIL (and no NPX is calculated) if there are < 10k read counts per sample, or if incubation, extension or amplification controls have <150 counts per sample. Blocks are flagged as FAILs (and no NPX values are calculated) if <3 plate controls or <1 negative control pass quality control. We excluded 19 samples due to 1) >50% of protein assays failed or 2) >50% of protein assays with counts below the average count in negative control samples.

#### SomaScan 11k v5

Proteomic profiling with the Somalogic SomaScan 11k v5 platform was performed in 1751 samples from 1555 individuals. Details of the SomaScan platform have been previously described in detail.^125^ Briefly, SomaScan relies on modified DNA-based aptamers that recognise their target protein. Aptamers are grouped across 3 dilutions bins: 20% (1:5), 0.5% (1:200) and 0.005% (1:20,000). Somalogic’s workflow includes the generation of scaling factors to account for intra- and inter-plate effects, for a hybridization normalisation, median signal normalisation, plate-scale normalisation and interplate calibration steps. Proteins are quantified with relative fluorescence units (RFUs) and finally normalised using Adaptive Normalisation by Maximum Likelihood (ANML). SomaLogic does not provide LOD values. We excluded 4 samples that were strong outliers with a median sample RFU more than 3 standard deviation away from the average of the population.

### Cross-platform protein target mapping and annotation

We extracted from each vendor UniProt identifiers^126^ for affinity targets that resulted in a list of 11,327 unique identifiers. For each unique UniProt identifier, we then mapped all platform-specific IDs that mentioned in their description the respective UniProt identifier to identify unique and overlapping targets across proteomic platforms. We subsequently used different databases^126–129^ to annotate protein coding genes, their genomic position (GRCh build 38), different protein characteristics, tissue- and cell-type expression patterns of protein coding genes, as well as predictions of secretome locations. Missing information on the genomic location of protein coding genes was manually obtained from GeneCards.^130^ A table summarising the protein target annotations across both platforms is outlined in **Supplementary Table 12.**

### Genotyping and imputation

Genotyping was performed using genomic DNA extracted from saliva samples obtained via Oragene saliva sampling kits. Individuals were genotyped on the Illumina GSA v3 chip + extra multi-disease content. Genotype calling and initial genetic data quality control was carried out using Illumina GenomeStudio version 2.0. Briefly, automated clustering using the GenTrain algorithm was performed using 1970 selected very high-quality samples at a subset of high-quality variants (autosomal variants in Hardy–Weinberg equilibrium with GenTrain scores >0.7, reflecting high-confidence clustering). Iterative rounds of manual and automatic reclustering were then performed to identify low-quality variants and samples, ultimately resulting in a dataset with >99% call rate at 637,829 SNPs. This cluster file was then applied to the remaining ∼50,000 samples to call genotypes. Samples were removed if they had lower call rates per sex than in the original batch (<99.2% for females, <99.5% for males). Of the 54,206 genotyped samples, individuals were removed due to a missing NHS number, discordant gender and genetic sex, implausible genetic duplicates (i.e. non-twin duplicates), resulting in a dataset of 51,176 individuals genotyped at 608,329 autosomal SNPs with a genotyping rate of >99.9%.

Following removal of rare variants (MAF < 0.0001), palindromic variants, and indels, genotypes were imputed to the TOPMED-r3 multi-ancestry imputation panel to genome build hg38 using the TOPMED imputation server ^131^. Following imputation we performed SNP quality control, filtering to common (MAF > 0.01) biallelic autosomal variants with <10% missingness, imputation quality (INFO score) >0.7 with no significant (p-value<1×10^-15^) deviation from Hardy–Weinberg equilibrium. Variants with duplicate positions were removed. Imputed dosages outside of the ranges 0−0.1, 0.9 −1.1 or 1.9 −2.0 were set to missing. We removed individuals with >10% missing genotypes. Genetic duplicate samples were identified with KING^132^—10 pairs of probable identical twins were identified, and one of each pair removed. Using principal component analysis, we identified and excluded <10 ancestral outliers who did not cluster with South Asian ancestry reference samples from the Human Genome Diversity Project and 1000 Genomes project.^133^ Following a clustering-based procedure to estimate categorical ancestry groupings (Bangladeshi or Pakistani), a further 62 participants were excluded due to ambiguous ancestry. We further excluded participants with missing covariate (age and sex) information, and those not included in the proteomic experiment resulting in a final set of 1,411 and 1,535 participants for genome-wide association testing for Olink and Somalogic protein targets, respectively.

### Whole Exome Sequencing

The Broad Institute performed ’Standard Germline Exome v6’ using Twist exome capture reagents and Illumina 150bp PE Novaseq 6000 sequencing. BWA-MEM was used to map to the reference genome hg38 with ALT contigs to produce single individual gVCF and cram output files. Preprocessing and variant calling was performed using the Exome Germline Single Sample 3.0.0 pipeline^134^ using Picard 2.23.8^135^, GATK 4.2.2.0 HaplotypeCaller^136^, and Samtools 1.11^75^. The Broad Institute performed basic quality control statistics and delivered crams with >85% bases at >20X Twist bait target coverage. Chromosome Y and MT variants were not called, and chromosome X variants were called diploid for females and males.

Sample quality control (QC) was applied to remove those with <85% bases at >20X coverage in Gencode exons; with the contamination estimate freemix >0.03; self-stated gender that did not match biological sex inferred from exome data (and could not be reconciled); without a valid NHS number, sample duplicates (the lowest coverage sample(s) were removed). Joint genotype calling was performed on these samples using HAIL and GATK GenotypeGVCFs using the Broad Institute Joint Genotyping pipeline.^134^ WES crams were compared to 44,396 Illumina GSAv3 chip genotyping samples by using 3,596 common (MAF>0.001) SNPs that are in both WES and GSA hard called genotypes (without imputation) to identify high confidence matches. WES samples with mismatches were removed after comparing them to the GSA data as likely recruitment or laboratory errors. We further removed samples not predicted to be of South Asian ancestry based on PCA and reference samples from the 1000 Genomes Project. Individuals were stratified into Bangladeshi, Pakistani and other South Asian groups based on PCA. Samples were further excluded if the number of SNVs, transition/transversion ratio, number of transitions, number of transversions, number of insertions, number of deletions, insertion/deletion ratio were outside the median ±6 median absolute deviations (MAD) compared to samples from the same population, or if the heterozygote/homozygote ratio, heterozygosity rate was higher than the median + 6 MADs (to avoid removing samples with high autozygosity).

A random forest model was trained on chromosome 20 using the true positive (high confidence variant sites discovered in the 1,000 Genomes Project, SNVs present on the Illumina Omni 2.5 genotyping array and found in the 1,000 Genomes Project ^133^, INDELs present in the Mills and Devine data^137^, HapMap3 ^138^ SNVs and INDELs) and false positive variants (QD<2 OR FS>60 OR MQ<30) as described above and then applied to the whole dataset. Features selected included quality by depth (QD), mean heterozygous allele balance, multiallelic site, strand odds ratio (SOR), mapping quality (MQ), number of alternative alleles at a site, variant type, site for which alleles include a ‘*’ allele, rank sum test for mapping qualities of reference versus alternative reads, multiallelic site containing SNVs and indels, rank sum test for relative positioning of reference versus alternative alleles within reads and allele type. Variants were ranked by their random forest score (i.e. the probability a variant reflects a true positive) and binned. We selected a random forest bin of 77 (true positive rate = 98.76%, false negative rate = 0.70%) and 57 (true positive rate = 95.65%, false negative rate = 4.88 %) to retain SNVs and indels, respectively.

Among variants passing variant QC criteria, we further tested a combination of random forest bin, depth (DP), genotype quality (GQ), heterozygous allele balance (hetAB) and call rate to perform genotype-level QC. Specifically for variants passing the given random forest bin, genotypes were set to missing if they did not pass one or more of the listed genotype QC criteria. To determine the optimal combination, we calculated the percentage of true and false positives, the transmitted / untransmitted synonymous singleton ration in trios (inferred using KING ^132^), the total number of Mendelian errors in trios and the mean number of heterozygous calls in runs of homozygosity (ROHs), for each combination of QC filters. For X chromosome variants, DP and GQ thresholds were tested separately for males and females and we calculated the mean number of heterozygous calls in non-pseudoautosomal regions in males instead of number of heterozygous calls in ROHs. The final set of filters selected were random forest bin 80, DP 10, GQ 20, hetAB 0.2 for SNPs, and random forest bin 80, DP 10, GQ 20, hetAB 0.2 and call rate 0.95 for SNVs; and random forest bin 44, DP 10, GQ 20, hetAB 0.3 and call rate 0.95 for indels. For subsequent rare variant genetic analyses, we retained participants with available Olink and Somalogic proteomic data that passed quality control.

### Genome-wide and exome-wide association analysis

#### Common variant (MAF > 1%) GWAS

We run genome-wide association testing for plasma levels of each of 5678 protein groups using the REGENIE software (v.4.0).^139^ For step 1, we used an LD-pruned (r^2^<0.2 or <0.9, respectively) set of common (MAF>1%) genotyped (GWAS) or sequenced (WES) markers with high coverage (missingness < 1%). For GWAS using GSA chip + TOPMED-r3 imputation, hard called genotypes from the GSA chip 51k sample data release were used, and filtered in plink2 using settings --geno 0.01 --indep-pairwise 500 50 0.2 --maf 0.05 and snpsonly and no XY chromosome data. This resulted in data on 103131 variants as input to REGENIE step 1. For exome sequencing using Twist exomes, final genotype calls after quality control from the 55k sample data release were used, and filtered in plink2 using settings --geno 0.01 --hwe 1e-15 --indep-pairwise 1000 100 0.9 --maf 0.01 and no XY chromosome data. This resulted in data on 78354 variants as input to REGENIE step1.

We included sex, age, age squared, inferred genetic ancestry (Bangladeshi or Pakistani ancestry), and the first 20 genetic principal components as covariates. In step 2, we run association testing again separately for imputed and sequenced genetic variants using the same set of covariates and default parameters for REGENIE.

#### Signal selection

Significant associations were selected for loci with regional clumping +/-500kb. We considered two significance thresholds 1) Bonferroni and 2) genome wide significance P< 5e-8. The identified signals across all proteins were then clumped by LD (R^2^>0.6) to identify sentinel variants across all protein targets. The most significant regional sentinel across proteins associated at a locus was selected as the sentinel variant at each locus.

#### Rare variant (MAF < 1%) ExWAS

Exome-wide analyses were conducted for all high-quality variants with a MAC >=5 using Regenie.^139^ Age, age squared, sex, inferred genetic ancestry (Bangladeshi or Pakistani ancestry), and exonic PCs 1-20 were used as covariates in both step 1 and 2. In step 2, we ran single-variant tests for variants with MAC>=5 and gene-based tests (burden, SKAT, and SKAT-O) for 3 variant consequence masks and considered an allele frequency cutoff <1%. Variant inclusion criteria for the masks defined were: 1) MASK_A – LOFTEE-HC-LOF, 2) MASK_B – MASK_A + deleterious missense (CADD > 20, polyphen > 0.446, SIFT Deleterious) and 3) MASK_C = MASK_A + MASK_B + other missense variants. Significant associations were considered at a Bonferroni significance threshold for each platform, as described above.

#### Variant annotation using VEP for WES analyses

Variants were annotated using Variant effect predictor (VEP) as previously described. The impact prioritised for a given variant using the pick prioritisation framework reported by VEP. Where more than one impact was prioritised for a given variant, for example where this variant maps to more than one gene, or non-coding region near multiple genes. The following was used to prioritise the annotation taken forward: 1) If one of these annotations is in cis with the targeted protein, use this annotation. This was done in two stages: 1.1) variant annotated to gene which encodes the protein target 1.2) variant within +/-500kb of the gene encoding the protein target. 2) if the variant is trans to the gene encoding the protein target, use max impact as specified by VEP to select the most severe impact for this variant HIGH, MODERATE, LOW, MODIFIER.

#### Novelty assignment – common variants (MAF≥1%)

For selected variant – protein association, we intersected the associated region (±500kb) with ones reported in 17 previous large-scale proteomic studies up to 30x larger than ours.^1,3,10–23^ If the region was not reported, we classified the pairing as ‘novel locus’. In case the region was reported, but for a different set of protein targets (based on UniProt), we classified the association as ‘novel protein at locus’ distinguishing between whether the protein was targeted by previous studies, or not.

#### Classification of artificial common pQTLs

We and others have previously provided evidence that some genetic variants are associated with measurement characteristics of assay used rather than genuine plasma proteins levels.^3,14,140^ We therefore implemented multiple analysis to flag such ‘artificial’ pQTLs. Firstly, we flagged all pQTLs associated with aptamers not targeting human proteins. Secondly, we used logistic regression models to test whether quality control metrics for proteins were significantly enriched for associations with protein targets. For example, whether the fraction of extreme protein measurements was particularly high/low for proteins associated with a given pQTL. We tested a total of 61 and 19 quality control measures provided or derived from SomaLogic and Olink, respectively. We finally, tested whether cis-pQTLs identified on either platform significantly modulated cross-platform correlations. To this end, we combined evidence from two statistical tests. We first tested for an interaction effect between the cis-pQTL on the linear association between Olink and SomaLogic measurements and then tested whether correlation coefficients within each genotype group differed significantly. The latter ensured that small differences in slope were not mistaken as differential correlation. Analyses were implemented using base R functions and the *cocor* package.

### Testing for non-additive effects

For each variant – protein pair, we tested for a potential departure from the additive genetic model by re-running association testing for this pair but introducing another term encoding heterozygous carriers into the model following previous work.^141^ Briefly, the additional term is a generic term to test for a significant departure from linearity across the three genotypes and we subsequently assigned potential models of inheritance based on computing log-likelihood ratio tests recoding the genotype according to three different models of inheritance based on the annotated reference allele. To distinguish between truly recessive/dominant effects and other deviations from normality, we additionally computed the gain in significance of either the dominant or additive model as suggested previously and kept only variant – protein association examples with a gain>1.^142^ These analyses were implemented in R v.4.3.1 using the REGNIE step 1 files to account for relatedness.

### Mitochondrial Genome Wide Association Analyses (mt-GWAS)

#### Mitochondrial DNA Variant Calling

Mitochondrial DNA variants can exist as heteroplasmies (variants that are in a portion of the mtDNA copies) or as homoplasmies (variants that are in all mtDNA copies). The proportion of copies of mtDNA that have a heteroplasmy is the variant allele frequency (VAF).

Mitochondrial DNA variants were called using a pipeline based off the one used by the gnomAD database.^143^ WES crams were filtered to only include unmapped reads or reads that were mapped to the mitochondrial genome. The reads were unaligned and converted to FastQ format using Picard^135^ RevertSam and SamToFastq. The unaligned reads were then mapped to the reference genome using BWA_MEM^144^ twice: first to the original version of the mitochondrial reference genome, and then to a version of the reference genome where the “cut point” was shifted by 8000bp. Because the mitochondrial genome is circular, it must be artificially linearized by choosing a location in the genome as an artificial cut point. By aligning the reads twice to different versions of the reference, this pipeline addresses the potential failure of reads that cross the cut point to align. Variants are then called from both the original and shifted alignments using gatk Mutect2.^145,146^ The positions of variants called from the shifted alignment are then corrected to match the original reference genome positions. The VCF created from the original reference genome is filtered to only include variants between 2001-14000 bp (variants that are not near the original cut point). The VCF created from the shifted reference genome is filtered to only include variants near the original reference’s cut point (1-2000 and 14001-16569). Variants at known artifact prone sites (301, 302, 310, 316, 3107, 10933, and 16182) and variants with a variant allele frequency (VAF) of less than 0.01 were also filtered out. The two VCFs for each individual were then concatenated into one VCF using bcftools^147^ concat. The VCFs of all individuals were merged using bcftools merge.

To facilitate the use of tools developed for the nuclear genome that require genotypes of 0/1/2, variants were converted to genotypes based on their VAFs as follows: <0.2 (homoplasmic reference) = 0, 0.2-0.8 (heteroplasmies) = 1, and >0.8 (homoplasmic for alt allele) = 2.

In addition to the removal of variants in artifact prone sites (identical to those removed by the gnomAD database), variants were removed entirely from the hard-coded file based on the following criteria: in known homoplasmy tracts (MT:300-317 or MT:16180-16193), indel stacks (variants where multiple indels occur in a given position), and common low-level heteroplasmies (variants that occur at a VAF of 0.01-0.5 in >0.1% of the population) following gnomAD^143^. Because of the germline bottleneck, heteroplasmies do not survive for many generations and are therefore not expected to be shared by distantly related individuals. Therefore, removing common low heteroplasmies can reduce spurious heteroplasmies that may be due to sequencing errors.

#### Association Testing

We conducted genetic association testing for all homoplasmic mitochondrial single nucleotide variants (mtSNVs) with a MAC > 5 and VAF >0.8 with all protein targets (Olink and Somalogic). The same analysis framework using Regenie and covariates described above for nuclear genome analysis was utilised to test associations of mtSNVs with protein levels using Regenie. Step1 files for common GWAS variants (See above) were used to adjust for relatedness and population structure. Significant associations were considered at a significance threshold of P < 5e-8. Variants were mapped to the mitochondrial genes based on the revised Cambridge Reference Sequence (GenBank number NC_012920.1), all positions are in human genome build 38, consistent with the nuclear genome results described above.

### Integration of disease associations

To integrate information on disease associations, we leveraged a comprehensive set of 191 custom disease endpoints using BI_PY^148^, 491 3-digit ICD10 codes, with at least 100 cases ascertained from the 43774 individuals with overlapping genetic and EHR data. Since binary traits were derived from ICD10 codes and custom code lists, there was partial overlap in phenotype definitions, each with varying sensitivity and specificity. Genetic associations with results from routine blood tests, as quantitative traits, were extracted and processed from EHR records using QUANT_PY.^149^

Genetic colocalisation analyses were performed using coloc^150^ for all genome wide significant (P < 5e-8) common cis-pQTLs where the lead pQTL variant was associated with a disease endpoint or quantitative trait at a threshold of p<10^-4^. A +/- 500kb region around the lead cis-pQTL variant was considered in downstream colocalisation analyses. Colocalisation was conducted using the coloc.abf() function using priors p_1_ =1e-4, p_2_ =1.e-4, p_12_=5e-6.

Evidence of a shared genetic signal, colocalisation, was considered at posterior probability H4 ≥ 0.7. Results were additionally filtered based on linkage disequilibrium between the lead variant for the cis-pQTL and corresponding disease/quantitative trait within the region (r^2^≥0.7). The latter ensured flagging potentially artificial colocalisation results due to mis-specified causal variant constellations and further that the cis-pQTL is the strongest outcome signal in the region, even if not reaching genome-wide statistical significance. A protocol, we have previously successfully established.^11,14^

We used the igraph R package^151^ to generate networks of protein-disease connections for these protein-disease pairs that had evidence of shared genetic signals. We further manually curated the custom disease outcomes and quantitative traits into the most relevant ICD10 chapter.

## ACKNOWLEDGEMENTS

Genes & Health is/has recently been core-funded by Wellcome (WT102627, WT210561), the Medical Research Council (UK) (M009017, MR/X009777/1, MR/X009920/1), Higher Education Funding Council for England Catalyst, Barts Charity (845/1796), Health Data Research UK (for London substantive site), and research delivery support from the NHS National Institute for Health Research Clinical Research Network (North Thames). We acknowledge the support of the National Institute for Health and Care Research Barts Biomedical Research Centre (NIHR203330); a delivery partnership of Barts Health NHS Trust, Queen Mary University of London, St George’s University Hospitals NHS Foundation Trust and St George’s University of London

Genes & Health is/has recently been funded by Alnylam Pharmaceuticals, Genomics PLC; and a Life Sciences Industry Consortium of AstraZeneca PLC, Bristol-Myers Squibb Company, GlaxoSmithKline Research and Development Limited, Maze Therapeutics Inc, Merck Sharp & Dohme LLC, Novo Nordisk A/S, Pfizer Inc, Takeda Development Centre Americas Inc.

We thank Social Action for Health, Centre of The Cell, members of our Community Advisory Group, and staff who have recruited and collected data from volunteers. We thank the NIHR National Biosample Centre (UK Biocentre), the Social Genetic & Developmental Psychiatry Centre (King’s College London), Wellcome Sanger Institute, and Broad Institute for sample processing, genotyping, sequencing and variant annotation. This work uses data provided by patients and collected by the NHS as part of their care and support. This research utilised Queen Mary University of London’s Apocrita HPC facility, supported by QMUL Research-IT, http://doi.org/10.5281/zenodo.438045

We thank: Barts Health NHS Trust, NHS Clinical Commissioning Groups (City and Hackney, Waltham Forest, Tower Hamlets, Newham, Redbridge, Havering, Barking and Dagenham), East London NHS Foundation Trust, Bradford Teaching Hospitals NHS Foundation Trust, Public Health England (especially David Wyllie), Discovery Data Service/Endeavour Health Charitable Trust (especially David Stables), Voror Health Technologies Ltd (especially Sophie Don), NHS England (for what was NHS Digital) - for GDPR-compliant data sharing backed by individual written informed consent.

Most of all we thank all of the volunteers participating in Genes & Health.

A favourable ethical opinion for the main Genes & Health research study was granted by NRES Committee London - South East (reference 14/LO/1240) on 16 Sept 2014. Queen Mary University of London is the Sponsor, and Data Controller.

AW was partially supported by the Friede Springer Cardiovascular Prevention Center at Charité - Universitätsmedizin Berlin, Germany.

## Author Contributions (CRediT Model)

Conceptualization: CL, DAvH

Data curation: JCZ, MK, AW, MP, DAvH, ESTB, AZ

Formal Analysis: AW, MP, JCZ, MZ, LK, DAvH

Resources: KAH, SF, DAvH, CL

Investigation: AW, AZ, ESTB, PFC, WN, MP, CL, DAvH

Funding Acquisition: DaVH, SF, CL

Supervision CL, DAvH, MP

Writing – original draft: AW, MP, CL

Writing – review and editing: all authors reviewed and commented on the manuscript prior to submission.

## Data Availability

Individual-level data from Genes & Health are available for bona fide researchers on application (https://www.genesandhealth.org/). Genome-wide summary statistics will be released upon publication.

## Code Availability

Associated code will be deposited on GitHub upon final publication.

## Supplementary Materials

Supplementary Note, including Supplementary Figures 1 and 2

Supplementary Tables 1 to 12

