## Supplementary Note for "Proteogenomic origins of disease in British South Asians"

**Cross platform comparison of common pQTLs for proteins measured by both Olink and Somalogic**

We observed weak-to-moderate correspondence (Spearman's r: cis-pQTL = 0.35; trans-pQTL = 0.09; **Supplementary Figure 1**) for a total of 1755 pQTLs passing filtering that were associated with proteins measured with both platforms. Low concordance was most likely explained by poor technical performance of at least one platform for a given protein (**Fig. 1d**) illustrating the need for cross-platform integration of pQTLs even for shared protein targets. In our three-platform technical comparison within the same individuals in Genes & Health.[6] we observed that replication of pQTLs identified for SomaLogic targets for the same project targeted by Olink is mostly about detectability in Olink, where as vice vera the lack of replication from Olink to Somalogic is mainly due technical incapability of the assays.

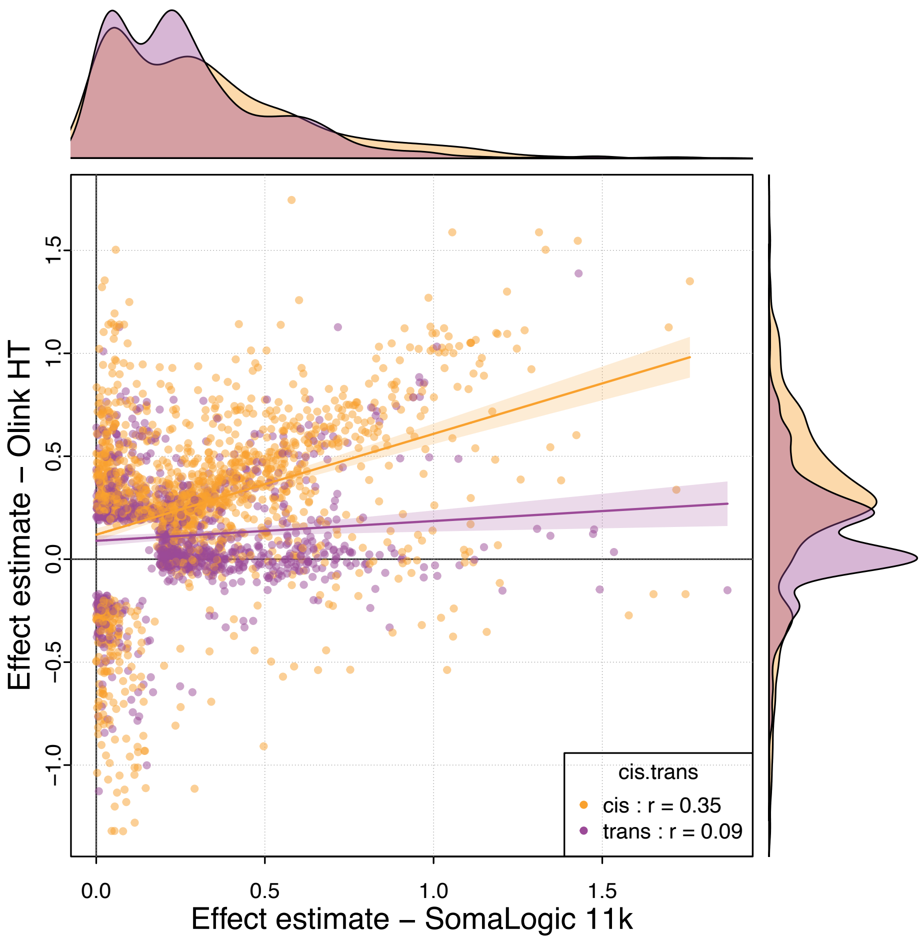


**Supplementary Figure 1: Comparison of effect sizes for cis and trans pQTLs for proteins measured by Olink and Somalogic.** Each dot is dot represents one of 1755 variant – protein associations that were filtered for artificial pQTLs and double counting (i.e, the same variant was identified for the same protein on the SomaLogic and Olink platform occurs only once). Associations are coloured by distance to the protein coding genes and correlation coefficients of effect sizes are given in the legend. Lines and associated 95%-confidence intervals indicate a linear fit.

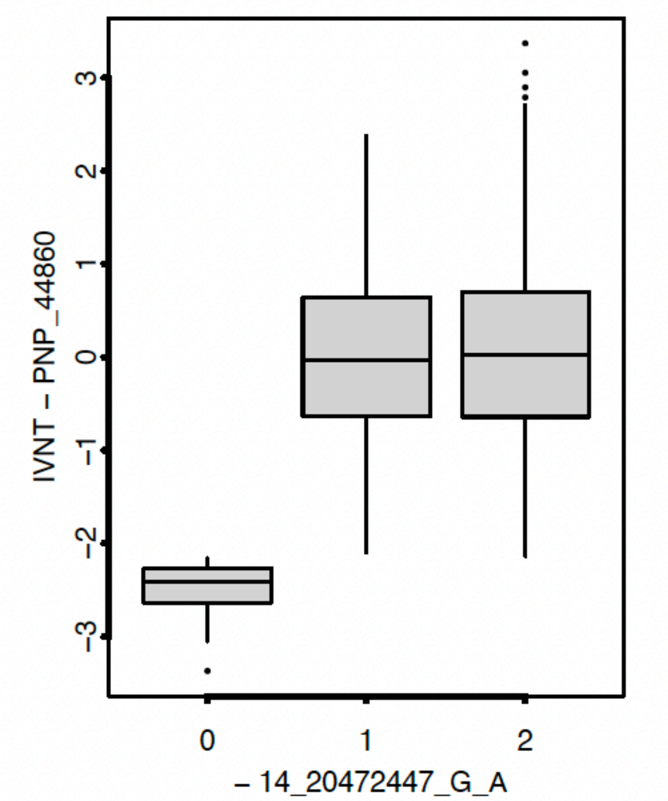


**Supplementary Figure 2: Non additive effects by genotype for the cis-pQTL for PNP**
x-axis denotes the genotype of carriers for the variant of interest – 0 – homozygous reference, 1 – heterozygous, 2 – homozygous alternate allele. Y axis denotes the inverse rank normalized protein abundance of PNP measured by the Olink platform (protein target - PNP_44860).
